# *Neisseria meningitidis* carriage, antimicrobial resistance, and risk factors in UK men who have sex with men

**DOI:** 10.64898/2025.12.03.25341543

**Authors:** Aminah Memon, Manik Kohli, Wenyu Liu, Emmi Suonpera, Yomna Gharib, Christos Karathanasis, Fowsiya Nur, Richard Gilson, Odile B Harrison

## Abstract

Urogenital infections caused by *Neisseria meningitidis* (Nm) are increasing globally, yet the prevalence, antimicrobial resistance profiles, and transmission dynamics of Nm in men who have sex with men (MSM) remain poorly defined. We conducted an oropharyngeal carriage study in 174 MSM attending a London sexual health clinic in 2023, prior to the implementation of 4CMenB vaccination and doxycycline post-exposure prophylaxis (doxyPEP). Nm was detected in 21.26% of participants, with carriage significantly associated with throat gonorrhoea, consistent with frequent co-colonisation. Whole-genome sequencing identified diverse lineages, including hyperinvasive clonal complexes (CC)11 and CC4821, and revealed widespread tetracycline resistance: 43% of isolates carried either a conjugative *tet(M)* plasmid or chromosomal *tet(B)* efflux locus. These findings indicate that MSM represent an important reservoir of tetracycline-resistant Nm, with the potential amplification of this phenotype following doxyPEP implementation, underscoring the need for genomic surveillance of *Neisseria* species in sexual networks.

## Introduction

*Neisseria meningitidis* (the meningococcus, Nm) is an opportunistic Gram-negative bacterial pathogen that spreads through aerosol transmission and colonises the human oropharynx. While the meningococcus generally exists as an asymptomatic commensal in approximately 10% of healthy individuals [1], invasive meningococcal disease (IMD) can manifest rapidly in infants, children, teenagers, and immunocompromised individuals and, more recently, has been increasingly observed in men-who-have-sex-with-men (MSM) [2].

Nm oropharyngeal carriage is a prerequisite for IMD, with transmission of Nm from carriers to susceptible individuals linked to an increased incidence of disease [3]. Typically, Nm carriage is associated with teenagers and adolescents due to their high carriage rates and increased risk for IMD [4–6]. High rates, however, have been recorded in MSM [7–9] with IMD outbreaks, predominantly driven by serogroup C Nm (MenC) belonging to clonal complex CC11, frequently reported in this population [10–13]. Social behaviours, including attendance at pubs/clubs, intimate kissing, and cigarette smoking are strongly associated with increased risk for meningococcal carriage in UK teenagers [4]. Similarly, sexual and social behaviours promoting frequent close contact will increase the risk for Nm transmission and carriage in sexually active groups including young adults, and MSM, with additional factors potentially reducing immunity such as HIV status [7].

In the UK, there is, however, limited information on Nm oropharyngeal carriage rates in MSM or how frequently Nm co-colonises with *Neisseria gonorrhoeae* (the gonococcus, Ng), a leading cause of sexually transmitted infection (STI), that has developed extensive antimicrobial resistance (AMR) [14, 15]. MSM frequently experience recurrent *N. gonorrhoeae* infections, including oral gonorrhoea. Combined with the high *N. meningitidis* carriage rates observed in this group, this suggests that co-colonisation of the oropharynx by both species is common, creating opportunities for interspecies horizontal gene transfer (HGT). Indeed, Nm and Ng isolates containing genetic determinants originating from either species have been documented consistent with HGT and leading to evolutionary adaptive changes that facilitate persistence in different anatomical niches, reduce susceptibility to antimicrobial treatment, or hinder detection using molecular diagnostics [16, 17].

To curb rising numbers of Ng cases reported in the UK, many of which increasingly antimicrobial resistant, the UK has introduced measures to prevent Ng infections, including vaccination with the Nm outer membrane vesicle vaccine (OMV), 4CMenB. This has been accompanied with the introduction of doxycycline post-exposure prophylaxis (doxyPEP) to limit other bacterial STIs with both offered to at-risk groups including MSM [18, 19]. Given the prevalence of Nm in MSM, these interventions will likely also affect Nm circulating in this population. We therefore undertook a cross-sectional, pilot study in MSM attending a sexual health clinic in central London, UK in 2023, pre-4CMenB and doxyPEP use, to assess oropharyngeal carriage and characterise the *Neisseria* population present. We detected a 21.26% Nm carriage rate with several serogroup B (MenB), tetracycline, azithromycin, ciprofloxacin, and penicillin resistant Nm found that had unknown predicted cross-reactivity with 4CMenB. The introduction of 4CMenB vaccination combined with doxyPEP therefore raises concerns on the selection and expansion of Nm exhibiting decreased antimicrobial susceptibility and indicate that further carriage studies are needed to understand *Neisseria* species dynamics, the incidence of Nm and Ng co-colonisation, and the potential impact of 4CMenB and doxyPEP on both Nm and Ng.

## Results

### Participant demographics, clinical NAATs results and *Neisseria* carriage rates

From May to September 2023, 174 MSM were recruited and provided oropharyngeal swabs from which 37 Gram negative, oxidase positive diplococci (GND+ve) were retrieved, all subsequently identified through whole genome sequencing as *N. meningitidis* (Nm) and resulting in a 21.26% (n = 37/174) carriage rate. Two samples were found to be mixed *N. meningitidis/N. gonorrhoeae* isolates.

No significant associations were found between Nm carriage and participant demographics (Table 1). Most participants were aged 25 to 54 with the highest number of participants in the 35-44 age range (55/174, 31.61%) closely followed by 25- to 34-year-olds (46/174, 26.44%) (Table 1). A total of 61 (35.06%) participants identified as white British with 48 (27.59%) identifying as white other background. Most participants reported living with one other person (86/174, 49.43%). The most frequent Nm carriers were aged 25 to 34 (12/37, 32.43%) with those aged 35 to 44 the most frequent non-Nm carriers (n = 47/137, 34.31%) (Table 1). Both carriers and non-carriers had similar proportions of partners (carriers: n = 35, 94.59%; non-carriers: n = 122, 89.05%).

**Table 1.**
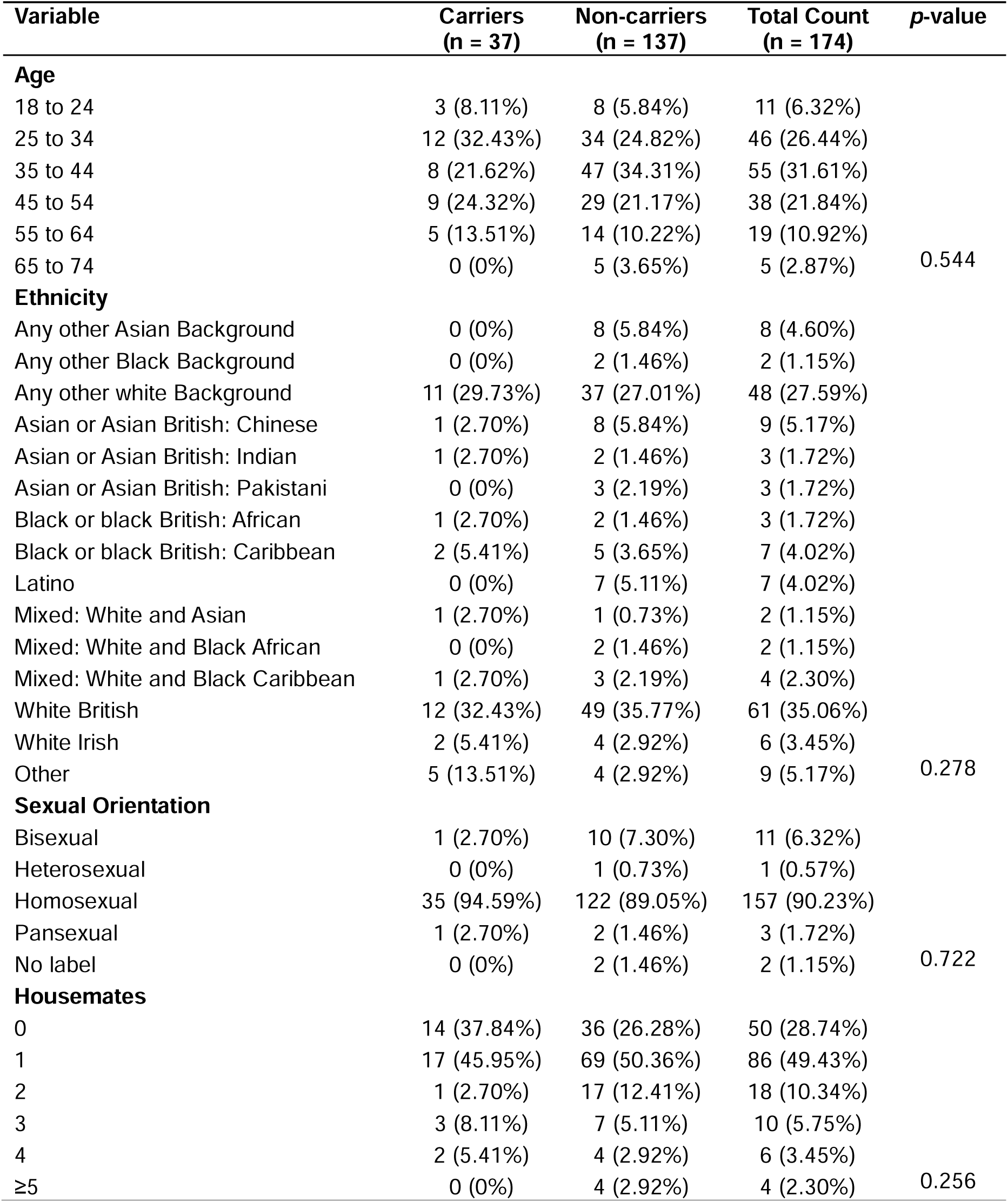
Participant demographics.

Clinical NAAT results from the 37 GND+ve positive samples, found seven (18.9%) positive for throat gonorrhoea, six (16.22%) for rectal gonorrhoea, and two (5.41%) for urethral gonorrhoea. One sample was positive for throat and urethral chlamydia respectively (2.70%), and two (5.41%) for rectal chlamydia. None were positive for syphilis while two participants reported having a cold/sore throat at the time of the study. Among Nm negative samples (n = 137), 10 (7.30%) were NAATs positive for throat gonorrhoea, 14 (10.22%) for rectal gonorrhoea, five (3.65%) for urethral gonorrhoea, four (2.92%) for throat chlamydia, eight (5.84%) for urethral chlamydia, and five (3.65%) for rectal chlamydia. Seven (5.12%) were positive for syphilis and 13 (9.49%) reported a cold/sore throat.

### Social, sexual, and clinical risk factors associated with Nm carriage

No causal variables were significantly associated with carriage after adjusting for confounders although social and sexual behaviours promoting close contact generally had higher odds of carriage (Figure 1). Smoking 11 to 20 cigarettes daily (aOR = 2.06, 95% CI: 0.40–8.73, *p* = 0.614) and waterpipe sharing (aOR = 4.18, 95% CI: 0.89–19.20, *p* = 0.186) experienced increased odds. Frequent sexual activity with multiple partners increased odds of carriage such as condomless anal sex three months prior to the study (aOR = 1.23, 95% CI: 0.44-4.02, *p*-value = 0.701) and having three or more male partners three months prior to the study (aOR = 1.96, 95% CI: 0.30-38.90, *p*-value = 0.295). Among non-causal exposures, a significant association between taking PrEP and carriage was found (aOR = 2.56, 95% CI: 1.03-6.78, *p*-value = 0.042) (Figure 2). Mutual masturbation (aOR = 1.90, 95% CI: 0.81-4.78, *p*-value = 0.140) experienced higher odds of carriage. Vaping, fisting someone, anal sex toy use, and getting fisted had aOR <1 in the adjusted models.

**Figure 1.**
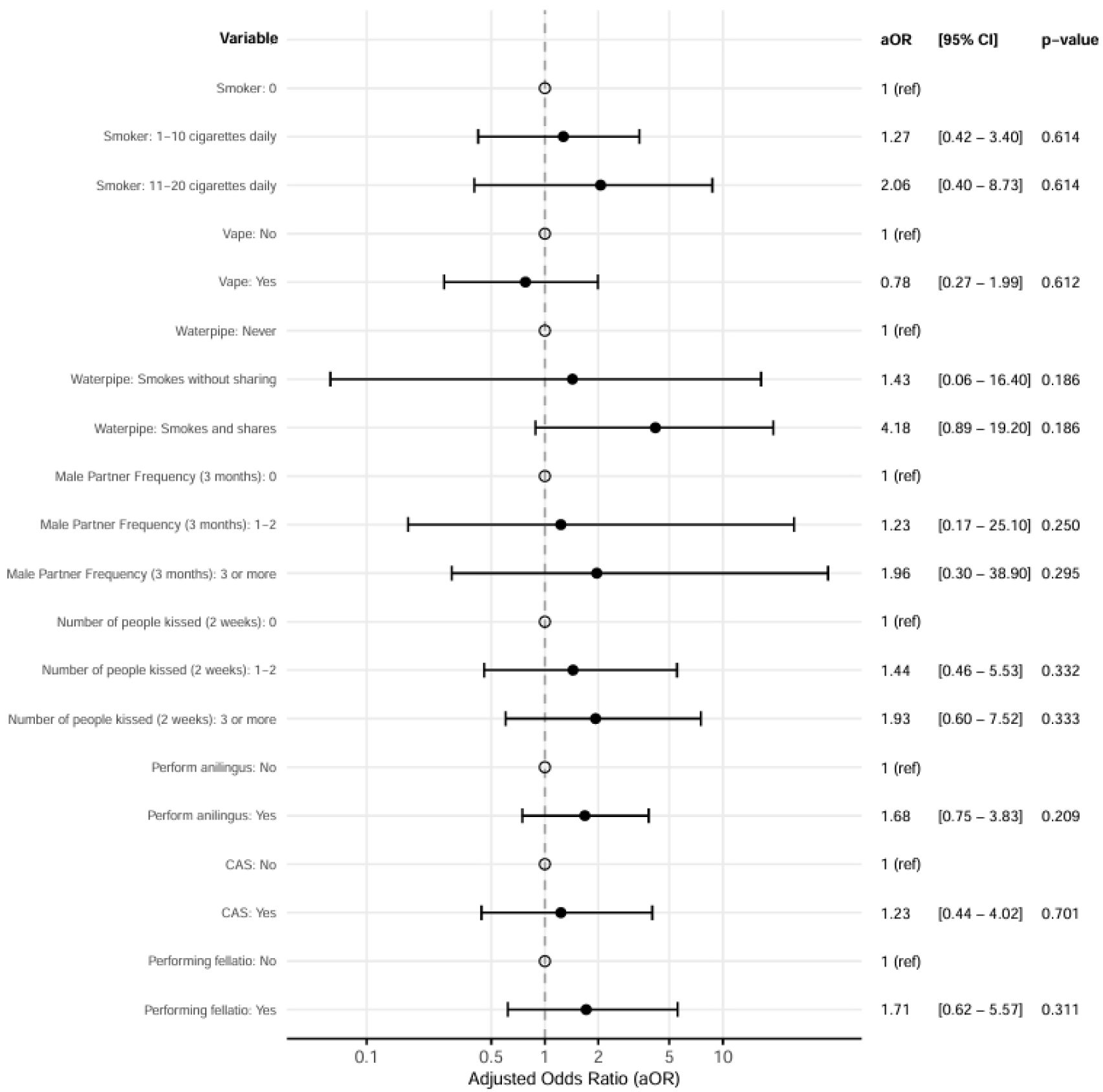
Forest plot depicting Behavioural adjusted odds ratios for causal exposures. 95% confidence intervals and *p*-values (derived from likelihood ratio tests) between behavioural and Nm carriage are depicted. Black-outlined circles represent reference values where OR = 1, black-filled circles represent adjusted odd ratios, and horizontal black lines represent 95% confidence intervals. The adjusted odd ratios were derived from multivariate logistic regression models for each causal exposure including covaries identified from the directed acyclic graph (DAG). aOR, adjusted odds ratio; CI, confidence intervals, Ref, reference value. *CAS: Condomless anal sex.

**Figure 2.**
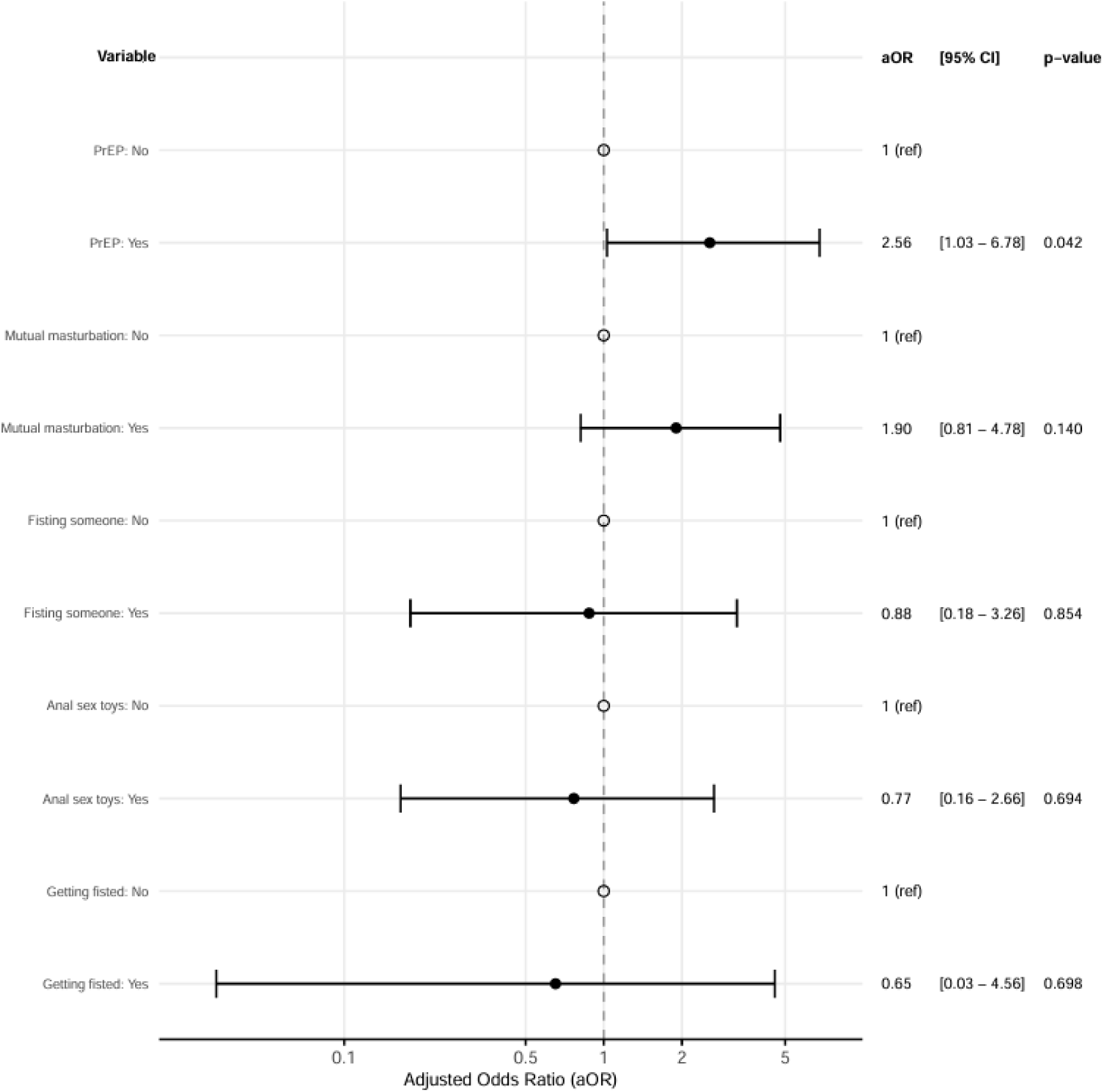
Forest plot depicting Behavioural adjusted odds ratios for non-causal exposures. 95% confidence intervals and *p*-values (derived from likelihood ratio tests) between behavioural and Nm carriage are depicted. Black-outlined circles represent reference values where OR = 1, black-filled circles represent adjusted odd ratios, and horizontal black lines represent 95% confidence intervals. The adjusted odd ratios were derived from multivariate logistic regression models for each non-causal exposure including covaries identified from the directed acyclic graph (DAG). aOR, adjusted odds ratio; CI, confidence intervals, Ref, reference value.

A positive throat gonorrhoea NAAT result was significantly associated with Nm carriage (OR = 2.96, 95% CI: 1.00-8.37, *p*-value = 0.049) with a positive urethral gonorrhoea NAAT result also significantly increasing odds of Nm carriage (OR = 5.11, 95% CI: 1.45-1.78, *p-*value = 0.012) (Table 2 & Figure 3). The throat and oropharynx are anatomical niches for both Nm and Ng, with higher odds of Nm carriage associated with positive NAATs results likely reflecting similar transmission pathways that are exploited by both pathogens, consistent with co-colonisation. Lower odds ratios were observed for chlamydia NAAT results in association with Nm carriage compared to gonorrhoea NAAT results, indicating that this was specific for gonorrhoea. Odds ratios <1 were observed for HIV positive, a positive rectal chlamydia NAAT result, and a cold/sore throat.

**Figure 3.**
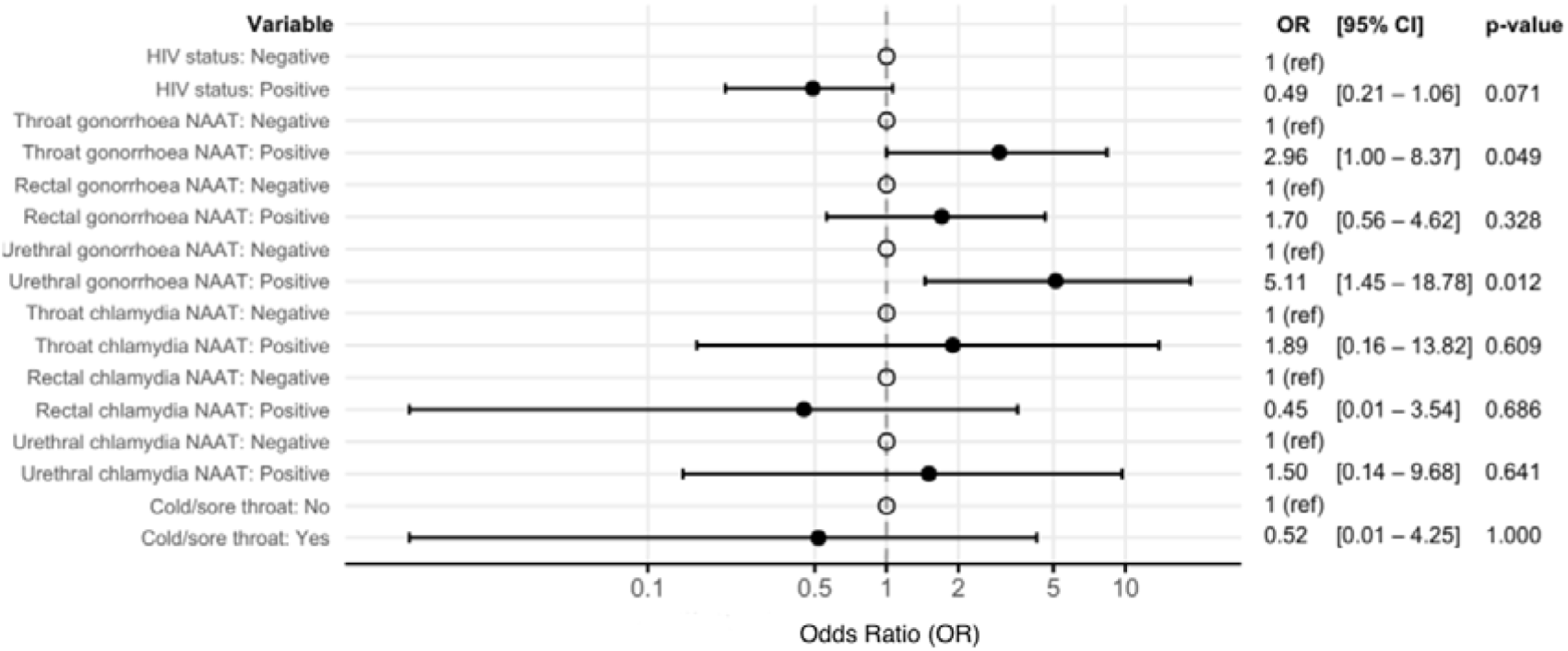
Forest Plot depicting adjusted odds ratios of clinical diagnoses and Nm carriage. 95% confidence intervals and *p*-values (derived from likelihood ratio tests) between clinical results and *Nme* carriage are depicted. ORs could not be calculated for syphilis due to zero responses in categories. Black-outlined circles represent reference values where OR = 1, black-filled circles represent adjusted odd ratios, and horizontal black lines represent 95% confidence intervals. OR, adjusted odds ratio; CI, confidence intervals, Ref, reference value.

**Table 2.**
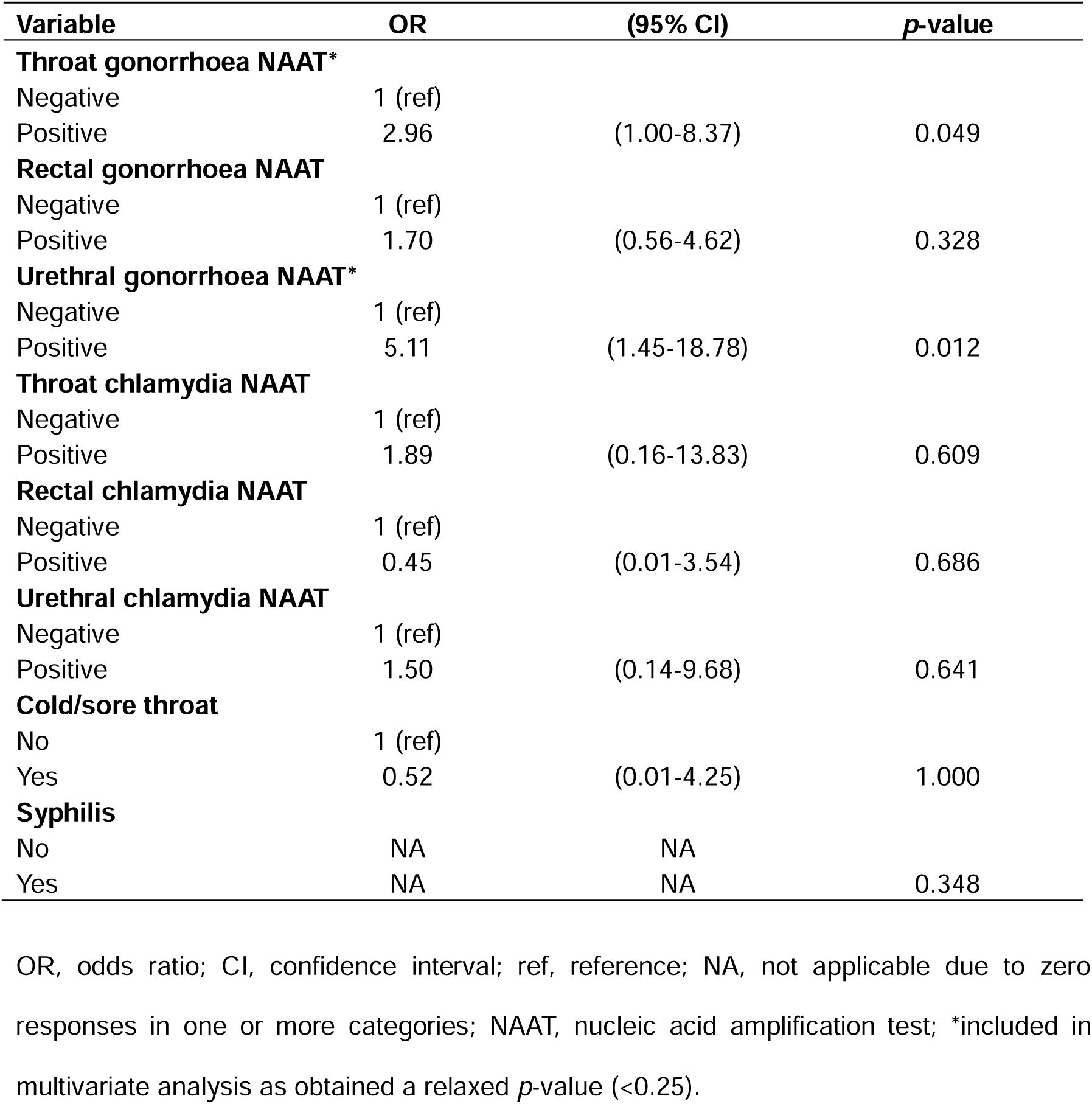
Univariate Analysis of Clinical Characteristics (n = 37)

| Variable | OR | (95% CI) | p-value |
| --- | --- | --- | --- |
| **Throat gonorrhoea NAAT\*** |  |  |  |
| Negative | 1 (ref) |  |  |
| Positive | 2.96 | (1.00-8.37) | 0.049 |
| **Rectal gonorrhoea NAAT** |  |  |  |
| Negative | 1 (ref) |  |  |
| Positive | 1.70 | (0.56-4.62) | 0.328 |
| **Urethral gonorrhoea NAAT\*** |  |  |  |
| Negative | 1 (ref) |  |  |
| Positive | 5.11 | (1.45-18.78) | 0.012 |
| **Throat chlamydia NAAT** |  |  |  |
| Negative | 1 (ref) |  |  |
| Positive | 1.89 | (0.16-13.83) | 0.609 |
| **Rectal chlamydia NAAT** |  |  |  |
| Negative | 1 (ref) |  |  |
| Positive | 0.45 | (0.01-3.54) | 0.686 |
| **Urethral chlamydia NAAT** |  |  |  |
| Negative | 1 (ref) |  |  |
| Positive | 1.50 | (0.14-9.68) | 0.641 |
| **Cold/sore throat** |  |  |  |
| No | 1 (ref) |  |  |
| Yes | 0.52 | (0.01-4.25) | 1.000 |
| **Syphilis** |  |  |  |
| No | NA | NA |  |
| Yes | NA | NA | 0.348 |

OR, odds ratio; CI, confidence interval; ref, reference; NA, not applicable due to zero responses in one or more categories; NAAT, nucleic acid amplification test; \*included in multivariate analysis as obtained a relaxed *p*-value (<0.25).

### *N. meningitidis* Isolate Characterisation

Thirty-seven Gram negative diplococci were retrieved of which 35 were identified as Nm (35/37, 94.59%). The remaining two isolates were mixed cultures, consisting of Nm and Ng, indicative of co-colonisation and were not further analysed here.

A total of 24 MLST STs were identified including five novel STs (Table 3). The most prevalent were ST-11 (5/35, 14.28%) and ST-5662 (5/35, 14.28%). Three isolates were ST-11563 (3/35, 8.57%), two isolates were ST-11312 and ST-5953 each (2/35, 5.71%), and 19 isolates possessed unique STs, identified once in this study. Most isolates did not belong to a clonal complex (CC) (n = 25/35, 71.42%). Of the 12 isolates that did, CC11 (5/35, 14.28%), CC4821 (3/35, 8.57%), CC198 (1/35, 2.85%), CC162 (1/35, 2.85%), and CC1136 (1/35, 2.85%) were identified. MenB made up most isolates (12/35, 34.28%), followed by group Z (6/35, 17.14%), MenW (4/35, 11.42%), and group X (1/35, 2.85%). Examination of the ST-5662 MenB isolates revealed that ST-17927 (id: 150945) and ST-11563 (id: 150924) isolates belonged to the same genome lineage and shared five of the seven MLST alleles indicative of an ST-5662 clonal complex (Supplementary Figure 2A). Eleven isolates were capsule null (*cnl*) (11/35, 31.42%). Three isolates (IDs: 150946, 150959 and 150935) (3/35, 8.57%) could not be assigned a capsule group.

**Table 3.**
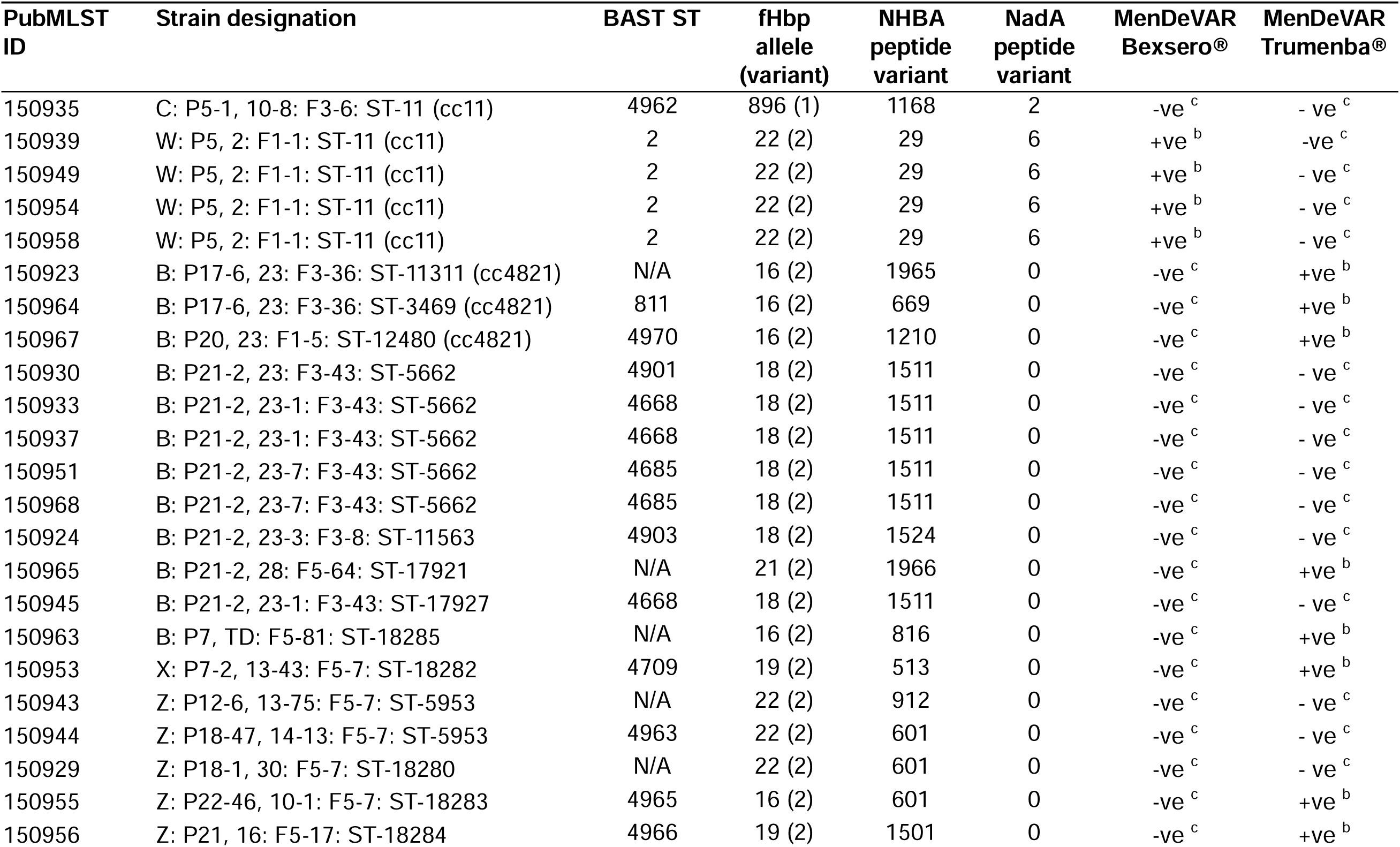
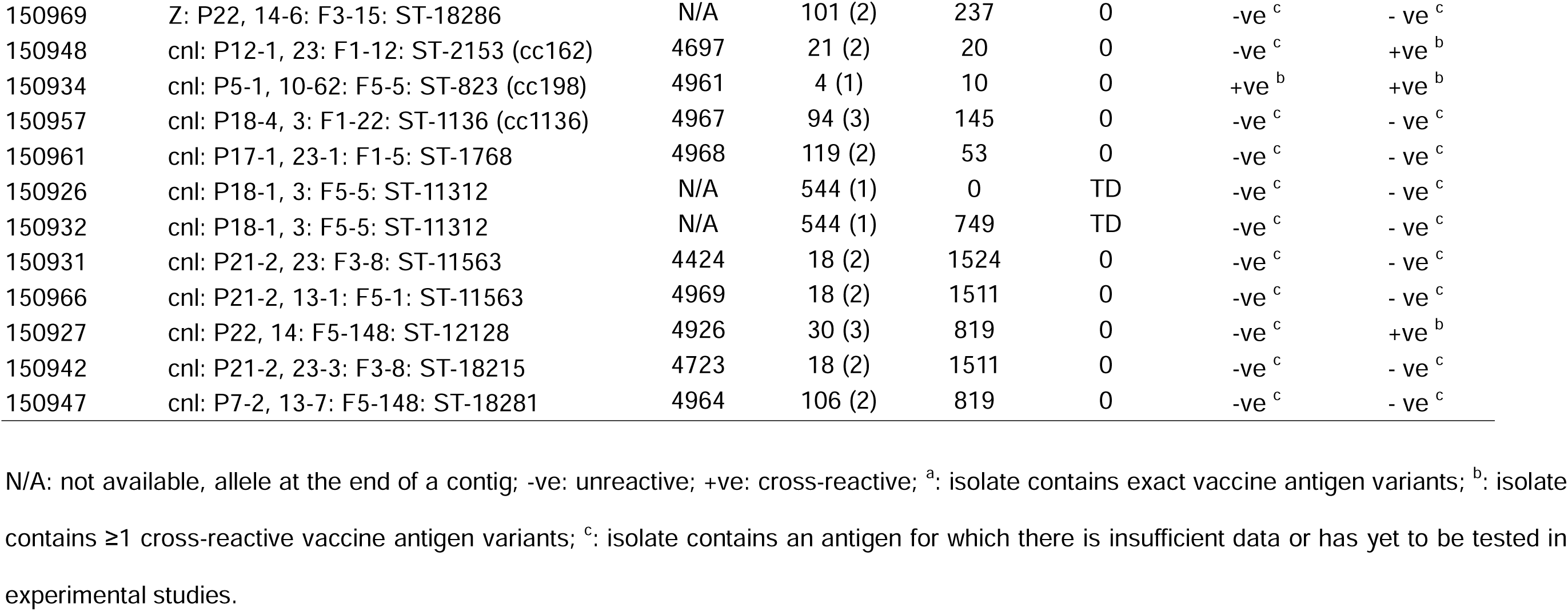
*N. meningitidis* isolate characterisation.

| PubMLST ID | Strain designation | BAST ST | fHbp allele (variant) | NHBA peptide variant | NadA peptide variant | MenDeVAR Bexsero® | MenDeVAR Trumenba® |
| --- | --- | --- | --- | --- | --- | --- | --- |
| 150935 | C: P5-1, 10-8: F3-6: ST-11 (cc11) | 4962 | 896 (1) | 1168 | 2 | -ve <sup>c</sup> | - ve <sup>c</sup> |
| 150939 | W: P5, 2: F1-1: ST-11 (cc11) | 2 | 22 (2) | 29 | 6 | +ve <sup>b</sup> | -ve <sup>c</sup> |
| 150949 | W: P5, 2: F1-1: ST-11 (cc11) | 2 | 22 (2) | 29 | 6 | +ve <sup>b</sup> | - ve <sup>c</sup> |
| 150954 | W: P5, 2: F1-1: ST-11 (cc11) | 2 | 22 (2) | 29 | 6 | +ve <sup>b</sup> | - ve <sup>c</sup> |
| 150958 | W: P5, 2: F1-1: ST-11 (cc11) | 2 | 22 (2) | 29 | 6 | +ve <sup>b</sup> | - ve <sup>c</sup> |
| 150923 | B: P17-6, 23: F3-36: ST-11311 (cc4821) | N/A | 16 (2) | 1965 | 0 | -ve <sup>c</sup> | +ve <sup>b</sup> |
| 150964 | B: P17-6, 23: F3-36: ST-3469 (cc4821) | 811 | 16 (2) | 669 | 0 | -ve <sup>c</sup> | +ve <sup>b</sup> |
| 150967 | B: P20, 23: F1-5: ST-12480 (cc4821) | 4970 | 16 (2) | 1210 | 0 | -ve <sup>c</sup> | +ve <sup>b</sup> |
| 150930 | B: P21-2, 23: F3-43: ST-5662 | 4901 | 18 (2) | 1511 | 0 | -ve <sup>c</sup> | - ve <sup>c</sup> |
| 150933 | B: P21-2, 23-1: F3-43: ST-5662 | 4668 | 18 (2) | 1511 | 0 | -ve <sup>c</sup> | - ve <sup>c</sup> |
| 150937 | B: P21-2, 23-1: F3-43: ST-5662 | 4668 | 18 (2) | 1511 | 0 | -ve <sup>c</sup> | - ve <sup>c</sup> |
| 150951 | B: P21-2, 23-7: F3-43: ST-5662 | 4685 | 18 (2) | 1511 | 0 | -ve <sup>c</sup> | - ve <sup>c</sup> |
| 150968 | B: P21-2, 23-7: F3-43: ST-5662 | 4685 | 18 (2) | 1511 | 0 | -ve <sup>c</sup> | - ve <sup>c</sup> |
| 150924 | B: P21-2, 23-3: F3-8: ST-11563 | 4903 | 18 (2) | 1524 | 0 | -ve <sup>c</sup> | - ve <sup>c</sup> |
| 150965 | B: P21-2, 28: F5-64: ST-17921 | N/A | 21 (2) | 1966 | 0 | -ve <sup>c</sup> | +ve <sup>b</sup> |
| 150945 | B: P21-2, 23-1: F3-43: ST-17927 | 4668 | 18 (2) | 1511 | 0 | -ve <sup>c</sup> | - ve <sup>c</sup> |
| 150963 | B: P7, TD: F5-81: ST-18285 | N/A | 16 (2) | 816 | 0 | -ve <sup>c</sup> | +ve <sup>b</sup> |
| 150953 | X: P7-2, 13-43: F5-7: ST-18282 | 4709 | 19 (2) | 513 | 0 | -ve <sup>c</sup> | +ve <sup>b</sup> |
| 150943 | Z: P12-6, 13-75: F5-7: ST-5953 | N/A | 22 (2) | 912 | 0 | -ve <sup>c</sup> | - ve <sup>c</sup> |
| 150944 | Z: P18-47, 14-13: F5-7: ST-5953 | 4963 | 22 (2) | 601 | 0 | -ve <sup>c</sup> | - ve <sup>c</sup> |
| 150929 | Z: P18-1, 30: F5-7: ST-18280 | N/A | 22 (2) | 601 | 0 | -ve <sup>c</sup> | - ve <sup>c</sup> |
| 150955 | Z: P22-46, 10-1: F5-7: ST-18283 | 4965 | 16 (2) | 601 | 0 | -ve <sup>c</sup> | +ve <sup>b</sup> |
| 150956 | Z: P21, 16: F5-17: ST-18284 | 4966 | 19 (2) | 1501 | 0 | -ve <sup>c</sup> | +ve <sup>b</sup> |
| 150969 | Z: P22, 14-6: F3-15: ST-18286 | N/A | 101 (2) | 237 | 0 | -ve <sup>c</sup> | - ve <sup>c</sup> |
| 150948 | cnl: P12-1, 23: F1-12: ST-2153 (cc162) | 4697 | 21 (2) | 20 | 0 | -ve <sup>c</sup> | +ve <sup>b</sup> |
| 150934 | cnl: P5-1, 10-62: F5-5: ST-823 (cc198) | 4961 | 4 (1) | 10 | 0 | +ve <sup>b</sup> | +ve <sup>b</sup> |
| 150957 | cnl: P18-4, 3: F1-22: ST-1136 (cc1136) | 4967 | 94 (3) | 145 | 0 | -ve <sup>c</sup> | - ve <sup>c</sup> |
| 150961 | cnl: P17-1, 23-1: F1-5: ST-1768 | 4968 | 119 (2) | 53 | 0 | -ve <sup>c</sup> | - ve <sup>c</sup> |
| 150926 | cnl: P18-1, 3: F5-5: ST-11312 | N/A | 544 (1) | 0 | TD | -ve <sup>c</sup> | - ve <sup>c</sup> |
| 150932 | cnl: P18-1, 3: F5-5: ST-11312 | N/A | 544 (1) | 749 | TD | -ve <sup>c</sup> | - ve <sup>c</sup> |
| 150931 | cnl: P21-2, 23: F3-8: ST-11563 | 4424 | 18 (2) | 1524 | 0 | -ve <sup>c</sup> | - ve <sup>c</sup> |
| 150966 | cnl: P21-2, 13-1: F5-1: ST-11563 | 4969 | 18 (2) | 1511 | 0 | -ve <sup>c</sup> | - ve <sup>c</sup> |
| 150927 | cnl: P22, 14: F5-148: ST-12128 | 4926 | 30 (3) | 819 | 0 | -ve <sup>c</sup> | +ve <sup>b</sup> |
| 150942 | cnl: P21-2, 23-3: F3-8: ST-18215 | 4723 | 18 (2) | 1511 | 0 | -ve <sup>c</sup> | - ve <sup>c</sup> |
| 150947 | cnl: P7-2, 13-7: F5-148: ST-18281 | 4964 | 106 (2) | 819 | 0 | -ve <sup>c</sup> | - ve <sup>c</sup> |

N/A: not available, allele at the end of a contig; -ve: unreactive; +ve: cross-reactive; <sup>a</sup>: isolate contains exact vaccine antigen variants; <sup>b</sup>: isolate contains $\geq 1$ cross-reactive vaccine antigen variants; <sup>c</sup>: isolate contains an antigen for which there is insufficient data or has yet to be tested in experimental studies.

The five CC11 meningococci were MenW (n = 4) with a non-groupable isolate (n = 1). The non-groupable isolate (150935, MSMCARR-045), NG: P1.5-1,10-8:F3-6:ST-11, was predicted to have a non-functional capsule due to the presence of the IS1301 insertion element in the capsule locus deleting the sialic acid biosynthesis genes *cssA*/*B*/*C* (NEIS0054-NEIS0052) and part of the capsule polymerase gene *csc* (NEIS0051). This isolate possessed alleles in the *norB-aniA* cassette (NEIS1546-NEIS1550), *ispD* (NEIS11438-NEIS1446), and arginine biosynthesis (*argB*) operon (NEIS1037-NEIS1038) that were identical to Nm from the urethritis clade, US_NmUC-A clade, first identified in the United States in 2015-2016 but observed globally since [20]. The four MenW isolates possessed the strain designation: W: P5,2: F1-1,ST-11 and belonged to the South American-United Kingdom MenW:CC11 sublineage responsible for MenW outbreaks in the United Kingdom and elsewhere beginning in 2009 and leading to the implementation in UK routine immunisation schedules in 2015 of the MenACWY polysaccharide-conjugate vaccine [21].

All Nm whole genome sequence data (WGS) were examined for antigen cross-reactivity with Bexsero (4CMenB) and Trumenba (rLP2086) using the MenDeVAR Index and the Bexsero antigen sequence typing (BAST) typing scheme [22, 23]. An *fHbp* gene was present in all except one Nm isolate. Variant 2 peptides were the most prevalent, found in 29/35 (82.85%) isolates with peptides 18 (n = 10), 22 (n = 7) and 19 (n = 2) accounting for 19/35 (54.28%) of the collection (Table 3). Variant 1 peptides were found in 4/35 (11.42%) of isolates with variant 3 found in two isolates. The *nhba* gene was present in all isolates with five Nm harbouring *nhba* genes predicted to be non-functional due to the presence of frameshift mutations resulting in premature stop codons. The *nadA* gene was absent in 30/35 (85.71%) isolates. A total of 22 BAST types were identified (Table 3). Of the 12 MenB isolates, none had sufficient data to predict 4CMenB vaccine coverage using available experimental data most likely due to fHbp and NHBA peptides for which no phenotypic data were available to allow predictions. Predicted coverage with rLP2086 was however observed for five MenB isolates, due to these possessing variant 2 fHbp peptides. Overall, four isolates (n = 5/35, 14.28%) were predicted to have antigens that were cross-reactive with 4CMenB, while 11 isolates were predicted to be cross-reactive with rLP2086 (n = 11/35, 31.42%).

### Genotypic and phenotypic antimicrobial resistance

#### Azithromycin

Phenotypic results showed that one isolate (1/35, 2.85%) exhibited medium-level resistance to azithromycin (16 mg/mL), six (6/35, 14.28%) low-level resistance (1-1.5 mg/mL) and seven (7/35, 22.85%) intermediate-level resistance (0.25-0.94 mg/mL), with the remaining 21 (21/35, 60%) isolates susceptible (0.047-0.19 mg/mL) (Table 4). None of the isolates exhibiting medium to low-level resistance (n = 13) possessed 23S rRNA mutations, however, they possessed MtrD alleles with the following amino acid substitutions known to confer resistance: (i) F854L (n = 1), (ii) K823E, F854L (n = 7), (iii) R714G; S821A; K823E; F854L (n = 1) and (iv) S821A; K823E; F854L (n = 2). Five of these isolates also possessed MtrR alleles with amino acid substitutions associated with resistance [24]. This included: (i) A39T; D79N (n = 1), (ii) D79N; A86T; S183N (n = 3), H105Y; S183N (n = 1). Three isolates possessed *mtrR* alleles lacking a stop codon. Allele 1216, found in one isolate (id 150923, B: P17-6, 23: F3-36: ST-11311 (cc4821)), harboured a premature stop codon, resulting in a predicted non-functional *mtrR*.

**Table 4.**
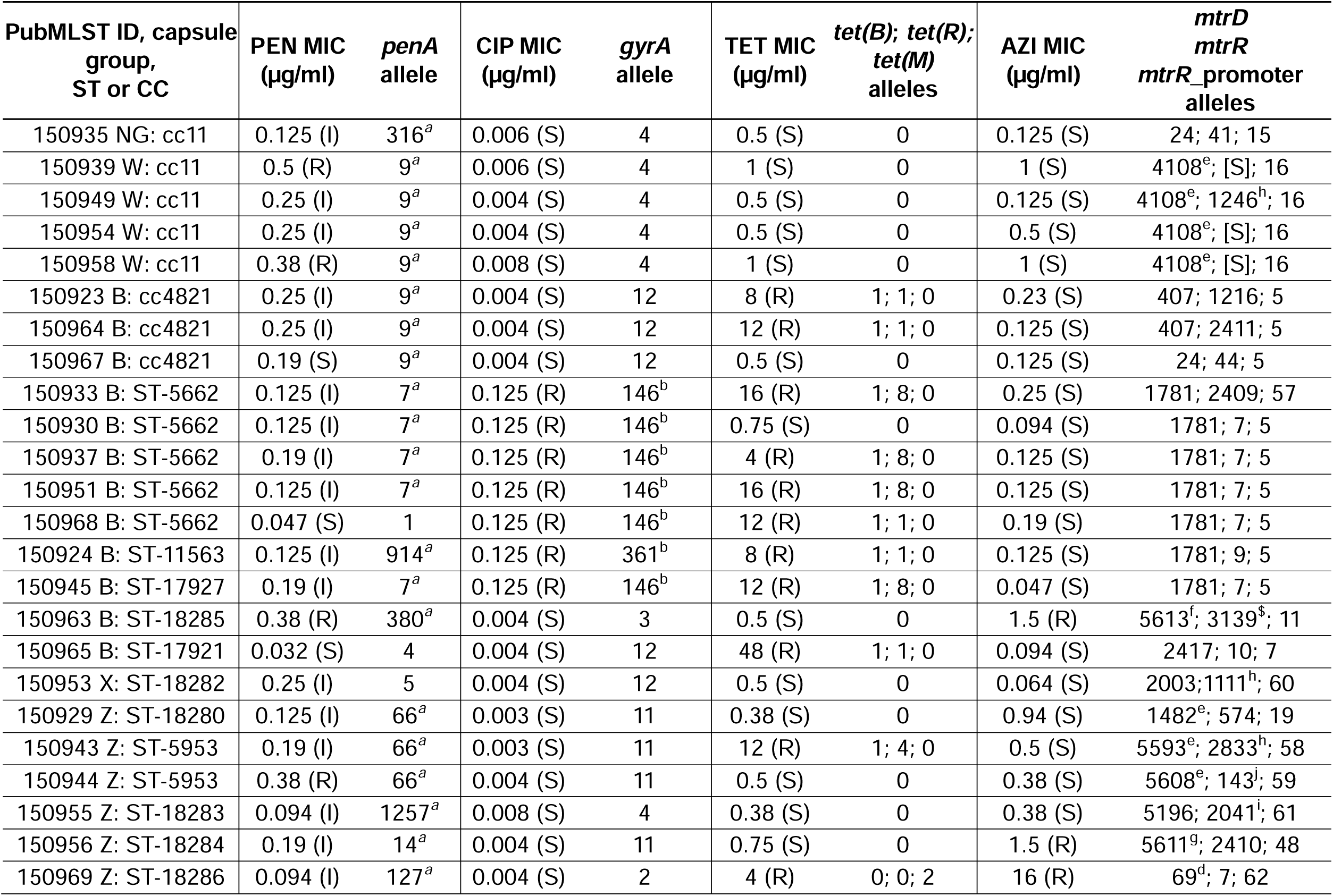
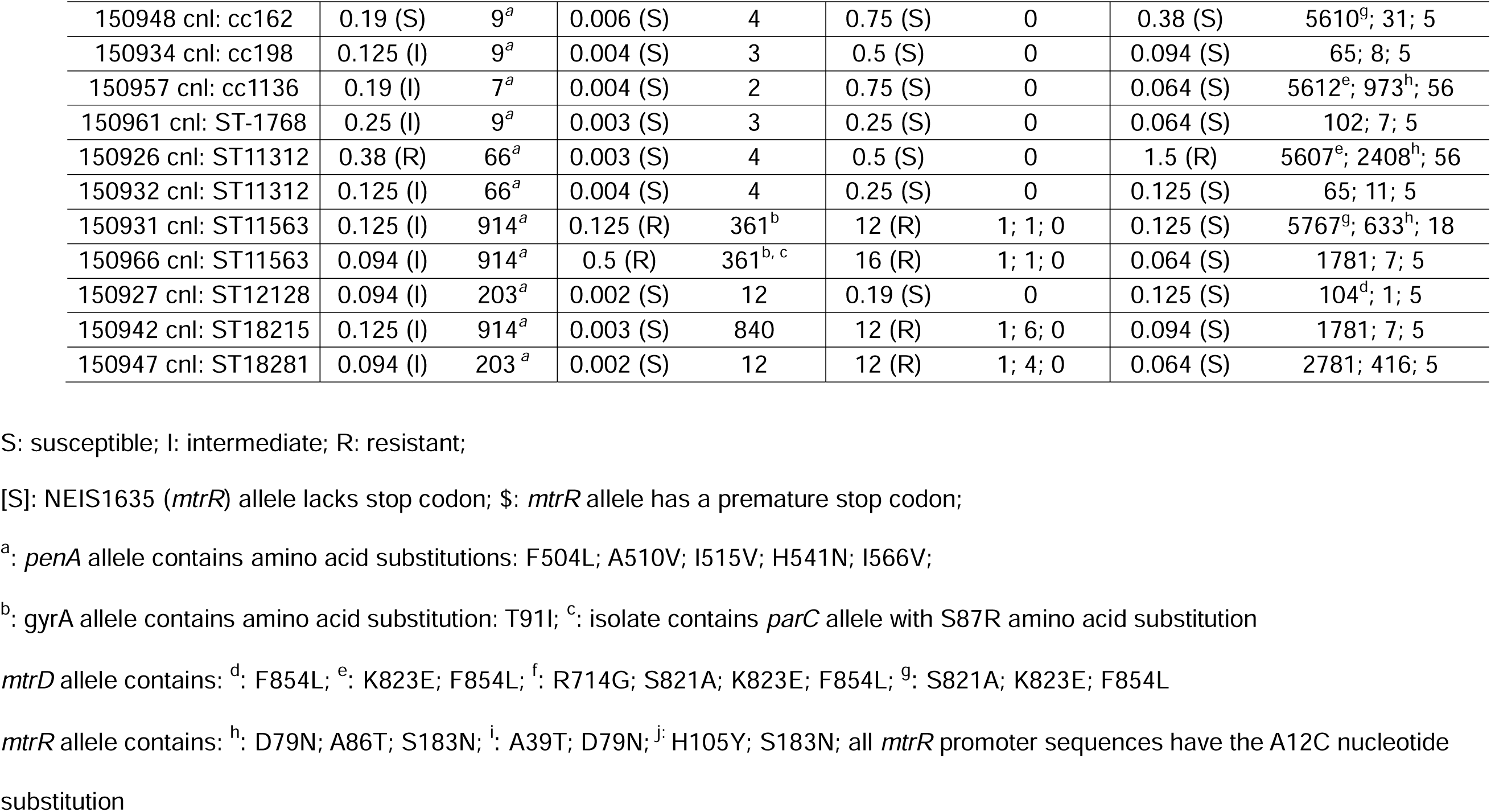
Isolate AMR genotypic and phenotypic results.

#### Penicillin

The majority of isolates (28/35, 80%) were categorised as penicillin-susceptible, increased exposure (PenI, 0.094-0.25 mg/mL). Two isolates were found to be penicillin-susceptible, standard exposure (PenS, MICs 0.032–0.047 mg/L). Five isolates were penicillin-resistant (PenR, 5/35, 14.28%, 0.38-0.5 mg/mL). The amino acid substitutions F504L, A510V, I515V, H541N, and I566V known to confer resistance to penicillin were identified in all PenI and PenR isolates (n = 33/35, 94.28%) and in 10 distinct *penA* alleles. No *penA* alleles were identified with the amino acid substitutions A311V, T483S, A501T, D511V, or G545S described in Ng and associated with reduced susceptibility to third-generation cephalosporins. The plasmid-borne beta-lactamase genes, *bla_ROB-1_* (NEIS3240) and *bla_TEM-1_*(NEIS2357), were absent in all isolates. Likewise, none of the *ponA* (NEIS0414) alleles contained the L421P amino acid mutation linked to penicillin resistance in Ng.

#### Ciprofloxacin

Nine (9/35, 25.71%; 0.125-0.5 mg/mL) isolates were resistant to ciprofloxacin. The remaining 24 isolates were susceptible (24/35, 68.57%; 0.002-0.008 mg/mL). Genotypic resistance was associated with the T91I substitution. None of the isolates harboured D95G/A/N/Y or T173N mutations in *gyrA* found in Ng. The amino acid substitution, S87R, in *parC* (NEIS1525) associated with increased ciprofloxacin resistance was identified in one isolate (n = 1/35, 2.85%), allele 243.

#### Tetracycline resistance

Fifteen isolates exhibited tetracycline resistance (15/35, 42.85%; 4-48 mg/mL). The remaining 20 isolates were categorised as susceptible (20/35, 57.14%; 0.19-1 mg/mL). One isolate (ID: 150969 Z: P22, 14-6: F3-15: ST-18286) carried the American conjugative plasmid (pConj) containing *tet*(*M*) (NEIS2210) allele 2 [25]. This allele shares 100% sequence similarity with Ng *tet(M*) alleles consistent with HGT [26]. A markerless pConj, lacking *tet*(*M*), was present in one isolate (ID: 150947, cnl: P7-2, 13-7: F5-148: ST-18281).

A total of 14 isolates (n = 14/35, 40%) possessed a chromosomally-encoded *tet*(*B*) locus, inserted between the catalase gene, *catA* (NEIS0211) and an RNA polymerase sigma factor (NEIS0212) (Figure 4). The locus consists of a *tet*(*B*) gene (NEIS2907) which functions as a tetracycline efflux pump and its repressor gene, *tetR*(*B*) (NEIS2927) [27]. All 14 isolates possessed the same *tet*(*B*) allele which demonstrated 100% sequence similarity with a *tet*(*B*) gene from *Haemophilus influenzae* (GenBank accession number CP131723). It also shared 99.92% identity with the complete nucleotide sequence of transposon Tn*10* (GenBank accession number AF162223). All *tet*(*B*)-positive isolates carried the tetracycline repressor gene *tetR*(*B*) (NEIS2927). Eleven of these isolates possessed functional *tetR*(*B*) genes with the remaining four *tetR*(*B*) sequences (allele 8) containing a premature stop codon resulting in an inactive repressor, consistent with constitutive expression of *tet*(*B*). Nine of the 14 *tetB* positive isolates were MenB (9/14, 64.28%), several belonging to the same lineages (Supplementary Figure 2B).

**Figure 4.**
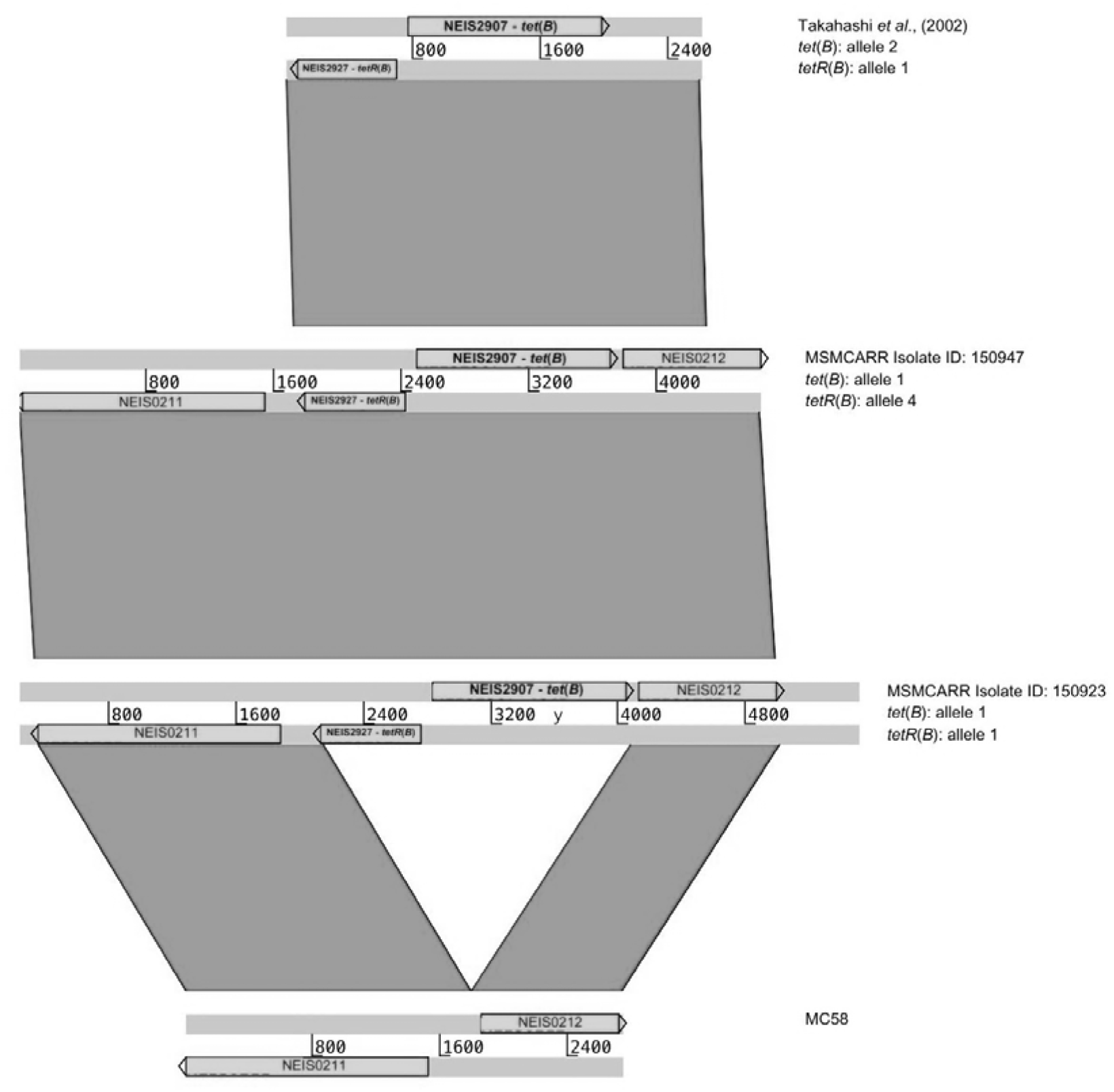
Artemis comparison of the *tet*(*B*) locus. Nucleotide Sequences were retrieved from the Takahashi *et al* study [27], accession number AB084246 and compared with the *tet*(*B*) locus from two MSM CARR isolates, 150947 and 150923. The *tet*(*B*) locus is flanked by the catalase gene, *katA* (NEIS0211) and an RNA polymerase sigma factor (NEIS0212). This region was extracted from the serogroup B reference genome, MC58 (accession number AE002098), which does not harbour TetB [67]. Comparisons were undertaken using the Artemis comparison tool (ACT) with the colour of the bands proportional to the percentage of sequence identity with the grey colour indicating 90 to 100% sequence identity.

We found a significant association between the presence of *tet*(*M*) and MSMCARR isolates. Of the 45,317 publicly available Nm whole genome records deposited in PubMLST, only 15 possessed a *tet*(*M*) pConj plasmid (MSMCARR: n = 1/35, PubMLST: n = 15/45,317; OR = 83.60, 95% CI: 1.94–579.26, *p* = 0.013). The same significant association was detected for *tet*(*B*) where of the 45,317 Nm WGS records, only 669 contained the *tet*(*B*) locus (MSMCARR: n = 14/35, PubMLST: n = 669/45,317; OR = 45.48, 95% CI: 21.84 - 92.47, *p* < 2.2 × 10^-16^).

### Concurrent AMR phenotypes

Tetracycline resistance (TetR) was the most frequent phenotype (n = 15), followed by reduced susceptibility to penicillin (PenR, n = 10), and ciprofloxacin (CipR, n = 9) (Figure 5). Nm with reduced susceptibility to azithromycin (AziReduced) was present in several isolates (n = 14). Co-resistance between tetracycline and penicillin (n = 2) or ciprofloxacin (n = 7) was observed, with only one isolate exhibiting triple-resistance combinations. No isolate exhibited concurrent resistance to all four agents tested. Overall, these data highlight the predominance of tetracycline resistance and the emergence of discrete clusters of dual or multidrug resistant meningococci that are circulating in the meningococcal population prior to doxyPEP and 4CMenB vaccination rollout.

**Figure 5.**
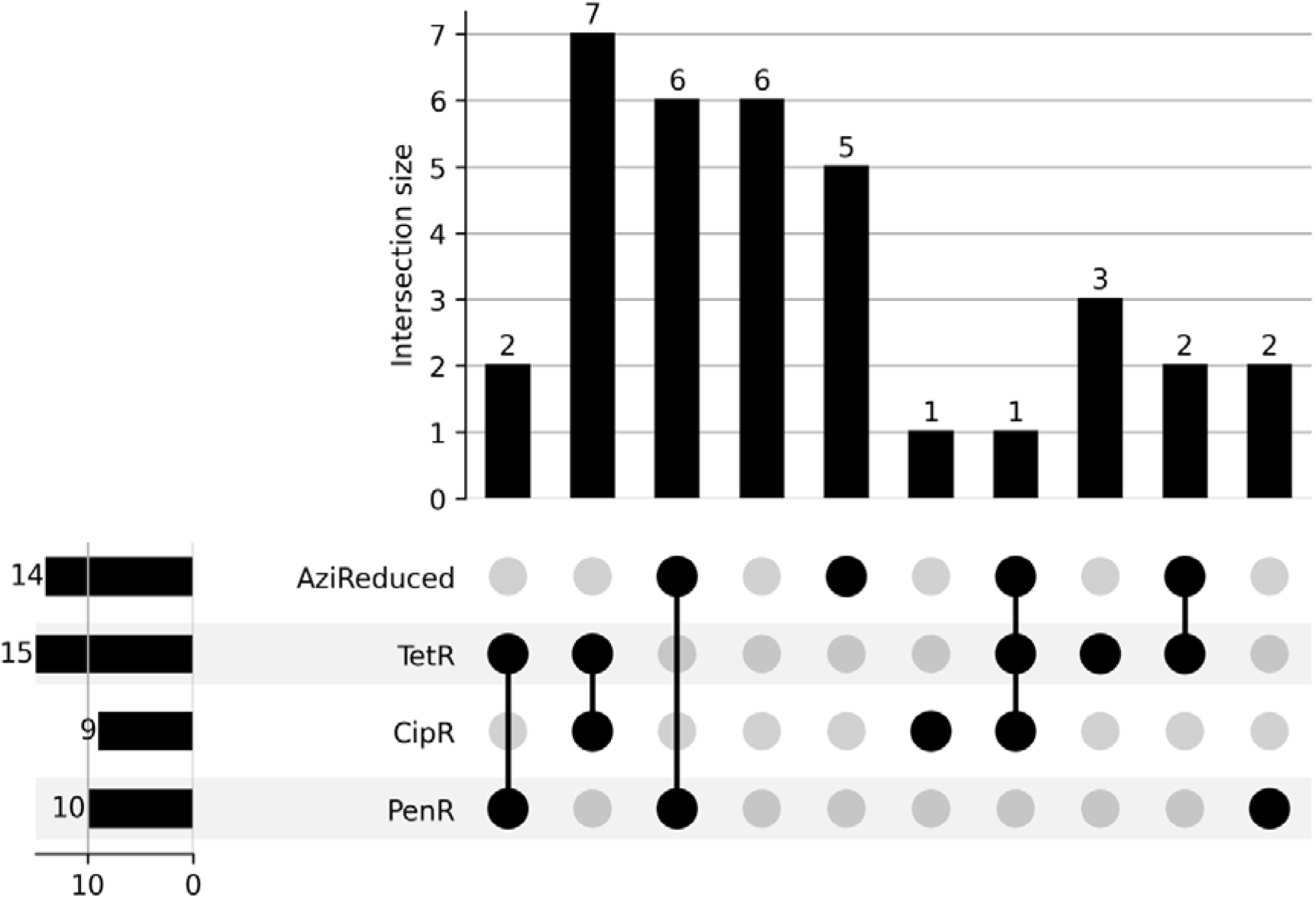
Upset plot of concurrent resistant phenotypes. Each bar represents the number of isolates sharing specific combinations of resistance phenotypes to penicillin (PenR), ciprofloxacin (CipR), tetracycline (TetR), and azithromycin (AziReduced). Four MIC categories indicative of reduced susceptibility to azithromycin were included when generating the Upset plot: intermediate (>0.25–0.5mg/mL), low-level resistance (1–2mg/mL) and medium-level resistance (4–32mg/mL). The TetR category included isolates with MICs in the 4-48 mg/mL range; CipR included isolates with MICs 0.125-0.5 mg/mL and PenR 0.094-0.5 mg/mL. Tetracycline resistance predominated (n = 15), with co-resistance found with penicillin (n = 2) or ciprofloxacin (n = 7) but only one isolate displaying triple-resistance combinations (AziReduced+TetR+CipR). Six isolates exhibited co-resistance with azithromycin and penicillin, No isolate displayed resistance to all four agents. These data indicate that tetracycline and azithromycin resistance is widespread in this dataset, and multidrug resistance is common.

## Discussion

*N. gonorrhoeae* (Ng) has developed extensive antimicrobial resistance (AMR), with both the number of cases and the proportion exhibiting resistance rising globally. In response, the UK has introduced preventative measures against gonococcal infections, including the *N. meningitidis* (Nm) vaccine 4CMenB [18, 28]. Although interventions such as doxyPEP and the 4CMenB vaccine aim to reduce bacterial sexually transmitted infections (STIs), their ecological impact on bystander *Neisseria* species remains poorly understood. In this study, meningococcal carriage was investigated in MSM before the implementation of 4CMenB and doxyPEP to establish a baseline for understanding how these measures may alter Nm carriage, antimicrobial resistance, and population structure.

Consistent with previous studies, we found a high Nm carriage rate (21.26%) in MSM, approximately fivefold greater than that reported in UK adolescents [8, 9, 29]. This elevated prevalence likely reflects behaviours promoting close physical contact and increased opportunities for oral-urogenital transmission. Over 90% of carriers reported recent oral or anal sexual contact, suggesting that Nm, like Ng, is transmitted within sexual networks and may colonise multiple anatomical sites. The significant association between Nm carriage and throat, urethral, or rectal gonorrhoea observed here supports frequent co-colonisation and shared transmission pathways between the two species.

In settings where oropharyngeal gonorrhoea and antibiotic use are common, co-carriage of *Neisseria* species facilitates interspecies recombination, accelerating the dissemination and selection of antimicrobial resistance determinants [30, 31]. The detection of Nm isolates with reduced susceptibility to penicillin (94%), ciprofloxacin (25%), tetracycline (42%), and azithromycin (37%) provides compelling evidence of this with the presence of both plasmid and chromosomally-mediated tetracycline resistance (*tet*(*M*) and *tet*(*B*) respectively suggesting multiple routes of AMR acquisition. The *tet*(*B*) locus shared near-identical sequence identity with *H. influenzae*, indicating horizontal gene transfer (HGT) across respiratory bacterial species. Such interspecies exchange likely occurs within the oropharynx, a densely colonised anatomical niche [32] where repeated antibiotic exposure, including doxycycline for STI treatment or prophylaxis, is likely to exert selection pressure. While azithromycin resistance in Nm is not yet widespread, cases have been recorded globally [33, 34] and, although IMD is not treated with azithromycin, this raises concerns for potential HGT to Ng, particularly given that MSM are at heighted risk of Ng infection.

The rarity of *tet*(*M*) and *tet*(*B*) in global Nm genome collections (0.03% and 1.5%, respectively, among >45,000 isolates) highlights the significance of this finding here. This disproportionate frequency indicates that antibiotic use within sexual networks may already be selecting for resistant Nm lineages prior to widespread doxyPEP implementation, possibly as a result of individuals self-sourcing antibiotics for STI prophylaxis [35]. The co-occurrence of multiple resistance mechanisms suggests that multidrug-resistant Nm populations are emerging through both HGT and local expansion. Future genomic analyses comparing mobile elements and resistance gene synteny between Ng, Nm, and commensal *Neisseria* could clarify the mechanisms underlying AMR gene flow.

The lower Nm carriage rates reported in adolescents compared to MSM may reflect differences in vaccine exposure. Introduction of the MenACWY conjugate vaccine in 2015 reduced MenW and MenC carriage through herd immunity [36, 37]. However, MSM are not routinely offered MenACWY, and our data show that the South American–UK MenW:CC11 sublineage is circulating in this group. The clinical significance of this finding is underscored by the atypical presentations and high fatality rates associated with MenW:CC11 IMD, highlighting the importance of continued vaccine awareness in sexual health settings [38].

Although the 4CMenB vaccine has reduced MenB IMD in the general population, its effect on Nm carriage is limited, likely due to antigenic diversity among circulating strains [39, 40]. The predominance of MenB isolates belonging to CC4821 and ST-5662 lineages, both harbouring *tet*(*B*) and exhibiting unknown predicted cross-reactivity with 4CMenB, suggests that these lineages may persist despite vaccination. These findings identify a genetically and epidemiologically distinct group of MenB meningococci associated with sexual transmission and exhibiting resistance to multiple antimicrobials, highlighting the need to monitor whether 4CMenB vaccination and HGT with Ng may further alter these lineages in MSM populations.

These results have broader public health implications. MSM experience disproportionately high antibiotic exposure due to recurrent STI treatment and emerging use of doxyPEP, creating an environment for AMR emergence across multiple commensal species [41]. Surveillance strategies should therefore include oropharyngeal *Neisseria* isolates from MSM, as resistance trends in this anatomical niche may precede those detected in Ng; the ‘canary in the coalmine’ effect [42]. Incorporating genomic AMR monitoring into UKHSA (UK Health Security Agency) sexual health frameworks could enable early identification of resistant meningococci or recombinant *Neisseria* species with clinical relevance. In line with the recent Chief Medical Officer’s 2025 Report [43] preventing future infections and unnecessary antimicrobial exposure is needed to slow the emergence and spread of resistance. Maintaining narrow-spectrum use of doxyPEP is necessary in slowing the spread of resistance, given that its use could indirectly influence STI transmission patterns through changes in perceived risk or prevention practices.

There are several limitations in this study. This was a single-centre, cross-sectional study with a small sample size and no longitudinal follow-up. Antibiotic exposure history was limited to the preceding two weeks, which may underestimate selection pressure. Furthermore, while the study excluded individuals recently treated with antibiotics, undetected subclinical exposures (e.g., post-treatment prophylaxis or unrecorded PrEP-associated antibiotics) may have influenced findings. Larger, multi-site, and longitudinal studies will be required to evaluate whether resistant Nm lineages expand following doxyPEP and 4CMenB implementation to determine how these interventions shape the oropharyngeal microbiome.

From an evolutionary perspective, the convergence of antibiotic pressure, high transmission, and multispecies colonisation creates an environment for genetic reassortment within *Neisseria* populations. MSM sexual networks thus represent important micro-ecosystems where AMR determinants may emerge and subsequently disseminate beyond the community through social and clinical contact. Understanding these processes is essential not only for STI control but also for anticipating resistance evolution in other *Neisseria* species.

In summary, this study identifies a high prevalence of Nm carriage and multidrug resistance in MSM prior to doxyPEP and 4CMenB rollout, including unprecedented rates of tetracycline and azithromycin resistance. These findings provide early evidence that sexual networks constitute a distinct environment for *Neisseria* evolution and AMR selection. As STI prevention strategies expand, integrated genomic and epidemiological surveillance will be critical to detect emerging resistant lineages, evaluate the impact of combined prophylactic interventions, and guide antimicrobial stewardship and vaccination policies in this at-risk population.

## Methods

### Participant recruitment and Isolate Collection

Adult (≥18 years) MSM attending the Mortimer Market Centre Sexual Health Clinic in London, United Kingdom between May to September 2023 were invited to take part in the study. MSM were defined as individuals who identified as men, including those assigned male at birth and transgender men. Charcoal-amies oropharyngeal swabs were collected regardless of gonorrhoea diagnosis and stored at 4lJ until plating. Participants who took antibiotics within two weeks prior to sampling were excluded. At the time of sampling, all participants completed a lifestyle questionnaire interrogating basic demographics, sexual behaviours, and clinical information with data stored on the REDCap database [44].

The study complies with relevant ethical regulations and was approved by the Health Research Authority and Research Ethics Committee (REC reference: 23/PR/0019). Written informed consent was obtained from participants. Participant’s samples and data were pseudonymised using a unique numerical identification number.

Nucleic Acid Amplification tests (NAATs) were conducted on self-collected Aptima® multitest swabs (Hologic Inc, Massachusetts, USA) using the Aptima® Combo 2™ assay on the Hologic Panther® system (Hologic Inc, Massachusetts, USA).

### Whole-Genome Sequencing and Isolate Characterisation

Within 24 hours of sampling, swabs were plated on chocolate-VCAT agar containing vitox with plates (Southern Group Laboratory) incubated at 37°C in a water-saturated atmosphere for 12 hours in 5% CO_2_. Gram-negative, oxidase-positive diplococci (GND +ve) were isolated and whole-genome sequenced using the Illumina MiSeq platform (Illumina, Inc., San Diego, USA). Raw sequence data were assembled using SPAdes [45] and resulting assemblies were uploaded to the genomics platform, PubMLST (https://pubmlst.org/neisseria) for species identification and isolate characterisation [46]. Annotations included capsule genogroup, MLST sequence type (ST), clonal complex (CC), fine-typing (PorA_VR1, PorA_VR2, and FetA_VR), and AMR loci including NEIS0414 (*ponA*), NEIS1753 (*penA*), NEIS1320 (*gyrA*), NEIS1525 (*parC*), NEIS2210 (*tet*(*M*)), NEIS2907 (*tet*(*B*)), and NEIS2927 (*tetR*(*B*)) (Table 5).

**Table 5.**
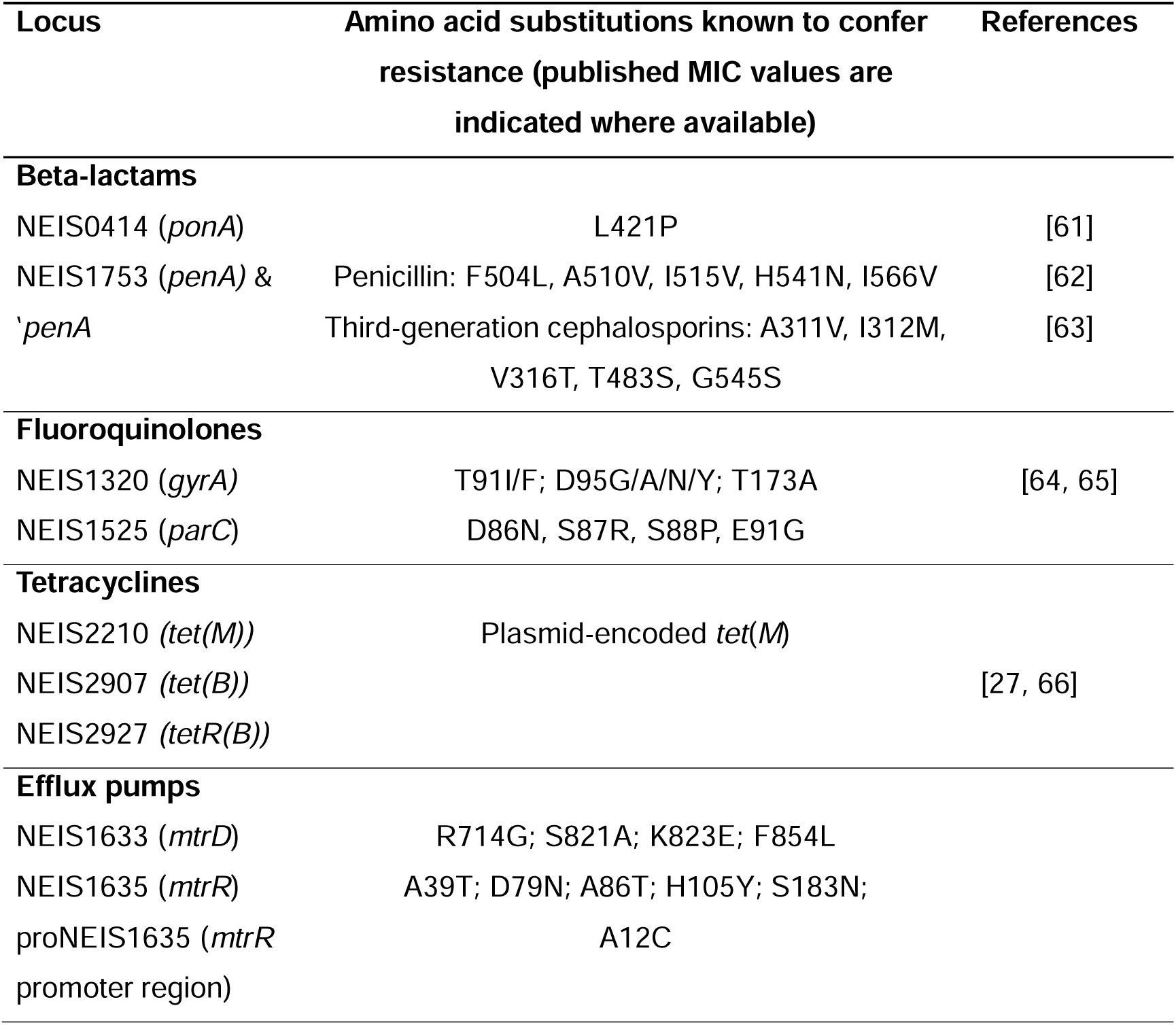
AMR Determinants analysed.

| Locus | Amino acid substitutions known to confer resistance (published MIC values are indicated where available) | References |
| --- | --- | --- |
| **Beta-lactams** |  |  |
| NEIS0414 (*ponA*) | L421P | [61] |
| NEIS1753 (*penA*) & 'penA | Penicillin: F504L, A510V, I515V, H541N, I566V<br>Third-generation cephalosporins: A311V, I312M, V316T, T483S, G545S | [62]<br>[63] |
| **Fluoroquinolones** |  |  |
| NEIS1320 (*gyrA*) | T91I/F; D95G/A/N/Y; T173A | [64, 65] |
| NEIS1525 (*parC*) | D86N, S87R, S88P, E91G |  |
| **Tetracyclines** |  |  |
| NEIS2210 (*tet(M)*) | Plasmid-encoded *tet(M)* | [27, 66] |
| NEIS2907 (*tet(B)*) |  |  |
| NEIS2927 (*tetR(B)*) |  |  |
| **Efflux pumps** |  |  |
| NEIS1633 (*mtrD*) | R714G; S821A; K823E; F854L |  |
| NEIS1635 (*mtrR*) | A39T; D79N; A86T; H105Y; S183N; |  |
| proNEIS1635 (*mtrR* promoter region) | A12C |  |

WGS data from 20,123 publicly available MenB isolates deposited in PubMLST were compared with MenB isolates from this study using the core-genome (cg) MLST scheme, cgMLST v3 [47]. A minimum spanning tree was generated and annotated by CC and *tet*(*B*) distribution.

### Predicted Vaccine Coverage

Prevention of IMD is possible either through immunisation using conjugate protein-polysaccharide vaccines targeting Nm expressing capsular serogroups A, C, W, and Y (e.g. MenACWY) or through the use of protein-based Nm vaccines including, 4CMenB (Bexsero; GSK) and rLP2086 (Trumenba; Pfizer, Inc), licensed for the prevention of MenB IMD. The protein antigens contained in these vaccines are different and display significant protein sequence diversity [48]. Both vaccines contain factor H binding protein (fHbp): one recombinant peptide variant in Bexsero (peptide 1) and 2 native lipidated peptide variants in Trumenba (peptides 45 and 55). Bexsero also contains the recombinant proteins neisserial heparin-binding antigen (NHBA [peptide 2]) and *Neisseria* adhesin A (NadA [peptide 8]) combined with the PorA-containing (variable region 2 [VR2]); peptide 4) outer-membrane vesicle from the MeNZB vaccine [22]. The diversity of 4CMenB vaccine antigens can be characterised from WGS using the Bexsero antigen sequence typing (BAST) scheme resulting in BAST sequence types which can be used to assess diversity and similarity to antigen variants included in vaccine formulations. The Deduced Vaccine Antigen Reactivity (MenDeVAR) Index [22] has in addition been developed to provide evidence-based information on the presence and possible immunological cross-reactivity of the different meningococcal vaccine antigen variants present in 4CMenB and rLP2086 and is publicly available on PubMLST. As the protein antigens in these vaccines are not MenB specific, all isolates were tested regardless of serogroup. The Bexsero^®^ Antigen Sequence Typing scheme [23] compares genomes with the Bexsero vaccine antigen variants: fHbp peptide 1, NHBA peptide 2, NadA peptide 8, and PorA VR2 4. The Trumenba^®^ reactivity analyses the fHbp peptide locus (peptides 45 and 55).

### Genotypic and phenotypic antimicrobial resistance

Loci defined in pubmlst.org/neisseria are allocated a value free nomenclature using the prefix NEIS followed by 4 digits. This includes a total of 10 loci commonly associated with AMR in Nm and Ng (Table 5). ResFinder v7.4.2 was used to screen for additional AMR genes that may not be defined in PubMLST, using the Basic Local Alignment Search Tool (BLAST) (http://blast.ncbi.nlm.nih.gov/Blast.cgi), searching for sequences with 80%-100% similarity [49]. Sequences identified using ResFinder were then queried against the database in PubMLST.org/neisseria to retrieve NEIS identifiers. Alleles designated by PubMLST for each isolate were downloaded as FASTA files and imported into MEGA11 [50] where deduced amino acid sequences were aligned using MUSCLE [51] and amino acid substitutions associated with AMR and internal stop codons were identified.

The *tet*(*B*) locus was confirmed by identifying: (i) an exact match to the NMB0217 open reading frame (ORF) (GenBank accession AE002379) from the Nm isolate, MC58, located 30 base pairs downstream of *tet*(*B*); (ii) a downstream ORF with 98% similarity to CP000381 from strain 053442 (GenBank accession CP000381.1) adjacent to *tetR*(*B*); and (iii) homology with Tn10 transposon. The *tet*(*B*) locus is flanked by the catalase gene, *catA* (NEIS0211) and an RNA polymerase sigma factor (NEIS0212). This region was extracted from the reference Nm strain MC58, two *tet*(*B*) positive Nm isolates from this study (Ids: 150947 and 150923) and the locus described by Takahashi *et al* [27]. Sequence data were then compared using the Artemis Comparison Tool [52].

Minimum inhibitory concentrations (MICs) for azithromycin, ciprofloxacin, benzylpenicillin, and tetracycline were determined using E-Tests (bioMerieux) and interpreted according to European Committee on Antimicrobial Susceptibility Testing (EUCAST) breakpoints v15.0 [53]. Antimicrobial susceptibility testing was performed on Mueller Hinton agar plates supplemented with 5% defibrinated sheep blood (Thermo Scientific Oxoid) and incubated at 37°C in 5% CO_2_.

Penicillin susceptibility was inferred as follows: isolates with penicillin MICs ≤0.06 mg/L were categorised as ‘susceptible, standard exposure (S)’ (where there is a high likelihood of therapeutic success using a standard dosing regimen of the agent) and isolates with penicillin MICs >0.25 mg/L were categorised as ‘resistant (R)’ (where there is a high likelihood of therapeutic failure even when there is increased exposure). Isolates with intermediate penicillin MICs were categorised as ‘susceptible, increased exposure (I)’ (where there is a high likelihood of therapeutic success because exposure to the agent is increased by adjusting the dosing regimen or by its concentration at the site of infection) [54]. Azithromycin breakpoints have not been established for Nm and therefore, four MIC categories were applied here based on azithromycin resistance described in Ng: susceptible (≤0.25 mg/L), intermediate (>0.25–0.5), low-level resistance (1–2), and medium-level resistance (4–32) [55].

The Upset plot was generated using a custom python script. Minimum inhibitory concentration (MIC) data for penicillin, ciprofloxacin, tetracycline, and azithromycin were converted into binary resistance variables based on EUCAST v15.0 clinical breakpoints. Values equal to or exceeding the resistance threshold were coded as *True* (resistant or reduced susceptibility), and all others as *False*. These datasets were then used to visualise resistance overlap patterns using an UpSet plot, generated in Python (v3.9) with the pandas, `matplotlib`, and `upsetplot` packages [56]. The `from_indicators` function was applied to aggregate intersection counts across all possible phenotype combinations, and the UpSet class was used to plot intersection sizes and shared resistance categories. All intersections with at least one isolate were displayed `(min_subset_size = 0)`, ensuring complete representation of resistance overlaps.

### Causal Inference Model and Statistical Analyses

Lifestyle questionnaire responses were used to construct a directed acyclic graph (DAG) in R version 4.3.2 (R Core Team, 2023) using `dagitty` [57], `ggdag` [58], `ggplot2` [59], and `DiagrammeR` [60]. The outcome variable was Nm oropharyngeal carriage (Supplementary Figure 1). Causal exposures were modelled as the following: HIV status, Perform anilingus, Condomless anal sex (CAS), Male partner frequency, Perform fellatio, Kissing frequency, Smoker, Vape, and Waterpipe. Non-causal exposures included Taken PrEP, Fisting someone, Getting fisted, Sex toys and Mutual masturbation. These variables and age were treated as confounders for other exposures. Multivariate logistic regression models for each exposure, including adjusted odd ratios (aOR), 95% confidence intervals (CI), and *p-values* were built based on the DAG. Confounders that were strongly correlated with the exposure were excluded from the model to avoid collinearity and overfitting. For example, in the exposure of ‘Kissing frequency’, the confounder ‘Male partners frequency’ was excluded as the behaviours were highly overlapping

Statistical tests were executed in R version 4.3.2 (R Core Team, 2023) using `dyplr, epitools`, `readxl,` and `tidyverse` packages [59]. Univariate logistic regression models were performed to analyse associations between clinical variables and carriage, and to obtain unadjusted odds ratios (OR). To prevent model overfit, categories with very low frequencies were combined with similar categories or excluded. The model was not run for demographic characteristics due to very low frequencies and high levels in each category. χ2 tests or Fisher’s exact tests were calculated where appropriate. ORs could not be calculated for factors with zero responses in categories. Fisher’s exact tests were conducted to assess whether the presence of *tet*(*M*) (MSMCARR: n = 1/37, PubMLST: n = 15/45,317) and *tet*(*B*) (MSMCARR: n = 15/37, PubMLST: n = 669/45,317), were more frequently associated with the Nm study isolates than with Nm WGS in PubMLST. Statistical significance was defined as *p*-values below 0.05 and recorded to three or more decimal places to prevent misinterpretation from premature rounding.

## Data availability

Whole genome sequence data generated in this study are available on the PubMLST database (https://pubmlst.org/neisseria).

## Author contributions

A.M. contributed data analyses, interpretation, and manuscript writing; W.L. provided statistical analyses; M.K. contributed study design, participant recruitment, consent and sampling, data analyses and manuscript editing; E.S. contributed study design, project management; Y.G. contributed participant recruitment, consent, sampling and manuscript editing; C.K. contributed participant recruitment, consent, sampling and manuscript editing; F.N. contributed participant recruitment, consent, sampling and manuscript editing; R.G. contributed manuscript editing and data interpretation; O.B.H contributed microbiology and molecular microbiology analyses, data analyses and interpretation, manuscript writing and editing.

---

**Supplementary Figure 1.**
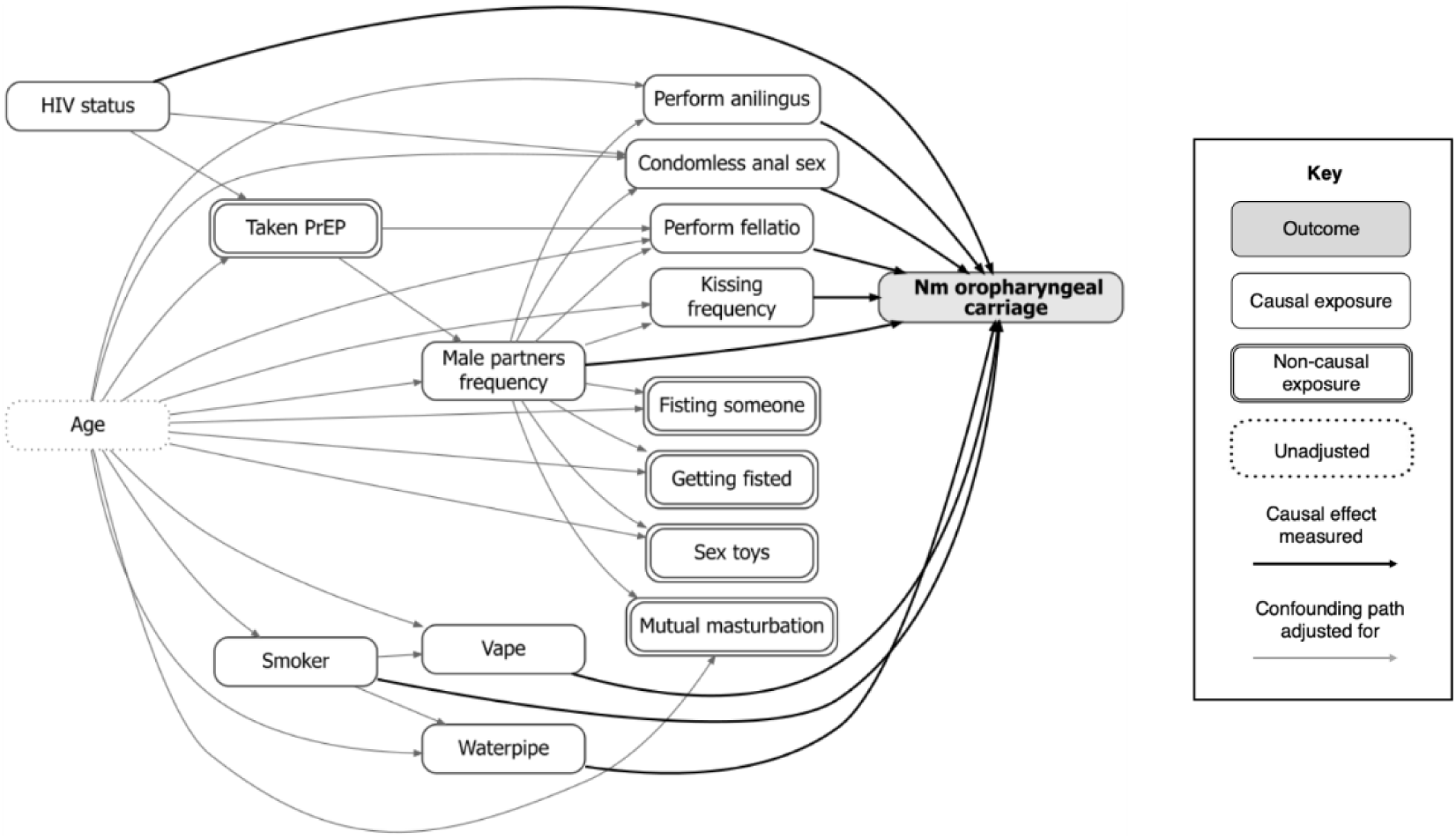
Directed acyclic graph (DAG) representing the variables from the participant lifestyle questionnaire estimated to have causal effects on Nm carriage.

**Supplementary Figure 2.**
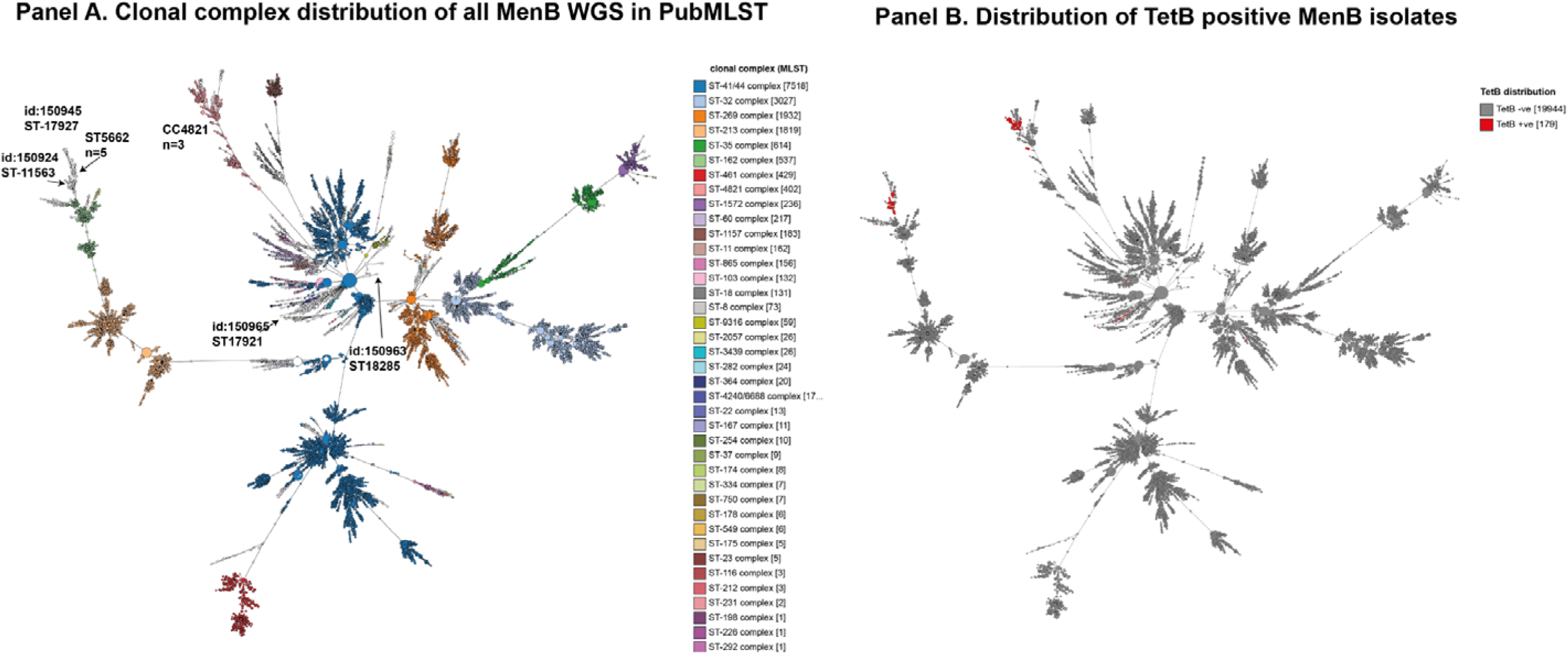
Comparison of MenB meningococci identified in this study with all MenB WGS in PubMLST. Whole genome sequence data from 20,123 MenB were compared using the cgMLST v3 tool in PubMLST after which a minimum spanning tree was generated. Each node represents an Nm strain which is coloured by clonal complex (Panel A) or TetB presence (Panel B). Clusters of isolates sharing allelic profiles in the core genome will form groups of related isolates. MenB isolates from this study are indicated in Panel A. This shows that the majority of isolates belong to 2 main lineages, CC4821 and ST-5662. Isolates from these lineages are also predominantly TetB positive.

